# Changes to household income in a Kenyan informal settlement during COVID-19

**DOI:** 10.1101/2021.06.15.21254693

**Authors:** Anna Tompsett, Aaron Baum, Vera Bukachi, Pascal Kipkemboi, Allan Ouko K’oyoo, Ana Varela Varela, Joseph Mulligan

**Affiliations:** Stockholm University, Stockholm, Sweden; Icahn School of Medicine at Mount Sinai, New York, USA; Kounkuey Design Initiative, Nairobi, Kenya; New York University, New York, USA; University of Amsterdam, Amsterdam, Netherlands; KTH Royal Institute of Technology, Stockholm, Sweden

## Abstract

More than a billion people live in densely-populated informal settlements worldwide. Crowded living conditions and limited resources may render these populations vulnerable to the health and economic effects of the COVID-19 pandemic. Representative and longitudinal survey data are needed to accurately measure impacts in these populations, but such data are scarce. Using satellite data and spatial sampling to ensure representativeness, we use longitudinal survey data on 1,033 households comprising 3,681 individuals collected pre- and post-pandemic to measure the economic effects of the COVID-19 pandemic households in six areas of the informal settlement of Kibera, Nairobi. The economic impacts are sizable and long-lasting. Household incomes declined at the start of the pandemic by 59% (95% CI: 50% to 69%) and remained 21% (95% CI: 13% to 28%) below the baseline level after six months. Respondents primarily attributed these declines in income to fewer labor market opportunities or lower demand for services, rather than the direct health impacts of the pandemic. The findings raise serious concerns about the welfare consequences of the COVID-19 pandemic for residents of informal settlements.

**Summary box:**

- Residents of informal settlements may be highly vulnerable to the economic impacts of the COVID-19 pandemic, but the longitudinal and representative survey data needed to measure impacts in these populations are scarce.
- We use an innovative spatial sampling strategy to collect representative survey data from residents in six areas of the informal settlement of Kibera, Nairobi.
- Relative to a pre-pandemic baseline, household incomes declined by 59% on average during the first three months of the pandemic. Six months later, incomes were still 21% below baseline.
- Households attributed declines in income primarily to fewer labour market opportunities or lower demand for services (43% of respondents) rather than the direct health impacts of the pandemic (1%).
- The declines in income we document raise concerns about the welfare of residents of informal settlements during the COVID-19 pandemic.

## Introduction

Worldwide, more than a billion people live in densely-populated informal settlements, of which just under a quarter live in sub-Saharan Africa (1). The well-being of these populations may be particularly vulnerable during the COVID-19 pandemic. Cramped living conditions and dependence on often-limited communal water and sanitation facilities increase vulnerability to infectious disease transmission, while many household’s incomes were insufficient to meet daily household needs even before the pandemic, implying a high level of vulnerability to interruptions in income (2). However, due to a lack of census data making it difficult to identify a representative sample and the high mobility of these populations, to date no representative longitudinal cohort studies have quantified the economic impacts of the COVID-19 pandemic on residents of informal settlements (3-9).

### Pre- and post-COVID-19 pandemic data for a representative sample of resident households in an informal settlement in Nairobi, Kenya

We analyzed data from a longitudinal survey of a representative cohort of 1,218 households (comprising 3,907 individuals) residing at six 100m-radius sites in the informal settlement of Kibera, Nairobi (**Figure 1a**). Kibera is Nairobi’s most populous informal settlement, located close to the city centre. The majority of dwellings are poorly-ventilated single room homes (10). Seroprevalence studies suggest that transmission in Kibera was likely to have been widespread during the study period (11, 12).

**Figure 1.**
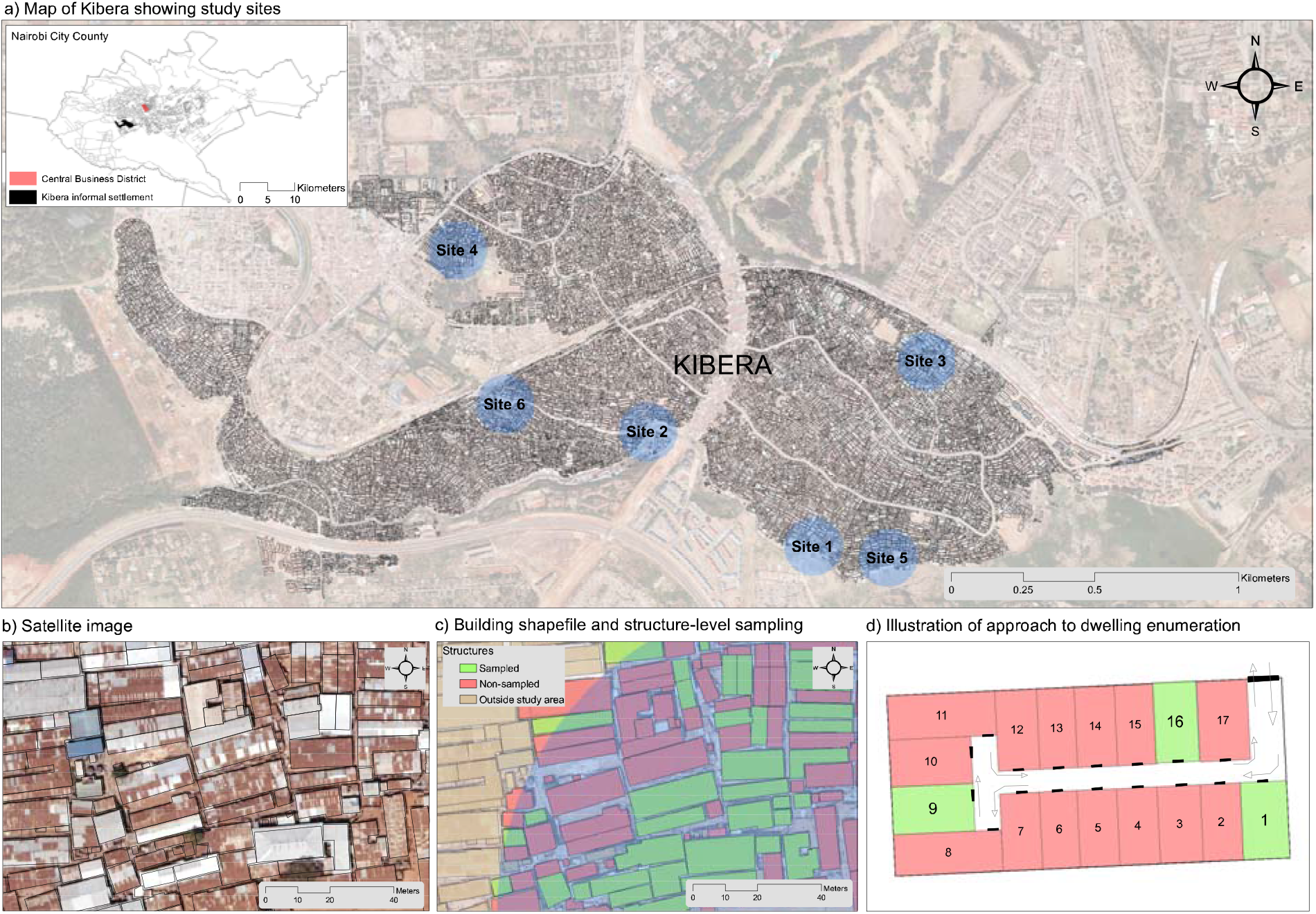
Map of study sites and sampling strategy a) Figure shows map of Kibera and location of each of six study sites within Kibera. Inset shows location of Kibera relative to Central Business District in Nairobi. b) High-resolution satellite imagery used to identify multi-household structures. c) Output from primary sampling process: i) structures outside study area shown in brown, sampled structures in green, non-sampled structures in red. d) Example illustration of door enumeration protocol. Enumerators turn to the left upon entering structure and number dwellings sequentially. Up to three dwellings randomly sampled from each multi-household structure.

The cohort was initially created prior to the pandemic to study the impacts of flooding in informal settlements. Given the lack of reliable population census data, we constructed a representative sample in two steps. First, we randomly sampled building structures from satellite images (**Figure 1b** and **1c**). Second, enumerators visited sampled structures to enumerate dwellings (**Figure 1d**), and we randomly sampled dwellings for interviews. After establishing a representative baseline sample, enumerators administered subsequent rounds of surveys approximately every six months by returning to the same dwellings and interviewing current residents. We use data from pre-COVID survey rounds that were conducted in-person between October 2019 and March 2020; during the pandemic, survey rounds were conducted by mobile phone between April 2020 and October 2020.

Respondents were asked to report their household income, whether that income was sufficient for their household’s needs, and reasons for changes to their income in all survey waves. Questions on respondents’ adoption of social distancing behaviors and self-reported health symptoms were added during the pandemic.

In the primary analysis, we restricted the sample to the subset of households (85% of 1,218 households) who were surveyed at least once before and once after the COVID-19 pandemic began in March 2020. We examined whether there were changes in mean household income between October 2019 and October 2020 using a linear regression model with adjustment for household fixed effects to account for pre-pandemic differences between households in income. Sampling weights were applied in order to account for stratification in the sampling strategy, and standard errors were clustered by the primary sampling unit (building structure). We examined the sensitivity of our results to potential bias from secular trends in mean household income, seasonal variation in household income, and non-random loss-to-follow up.

Analyses were performed in Stata v16 (StataCorp). Participation in the study was voluntary, and analyses were conducted on anonymized data. The Kenyan National Commission for Science, Technology & Innovation approved this study.

### Household incomes declined at the start of the pandemic and had not recovered after six months

The final sample consisted of 1,033 households comprising 3,681 individuals, with a median (IQR) age of 21 (10-33) and 49.8% female. Prior to the pandemic, enumerators completed a field interview with a respondent inhabitant in 87% of randomly sampled occupied dwellings. During the pandemic, enumerators successfully surveyed 85% of previous respondents.

Relative to the six-month period preceding the pandemic, self-reported weekly household income declined by 59% (95% CI: 50% to 69%) from 2324 KSh (US$21) to 1417 KSh (US$13) during April to July 2020 (**Figure 2**). By September 2020, self-reported household income partly rebounded but remained 21% (95% CI: 13% to 28%) below the baseline level. The fraction of households reporting income sufficient for their needs declined by 49% (95% CI: 34% to 64%) from 18% to 9% during April to July 2020, relative to the baseline period (**Supplemental Figure 1**).

**Figure 2.**
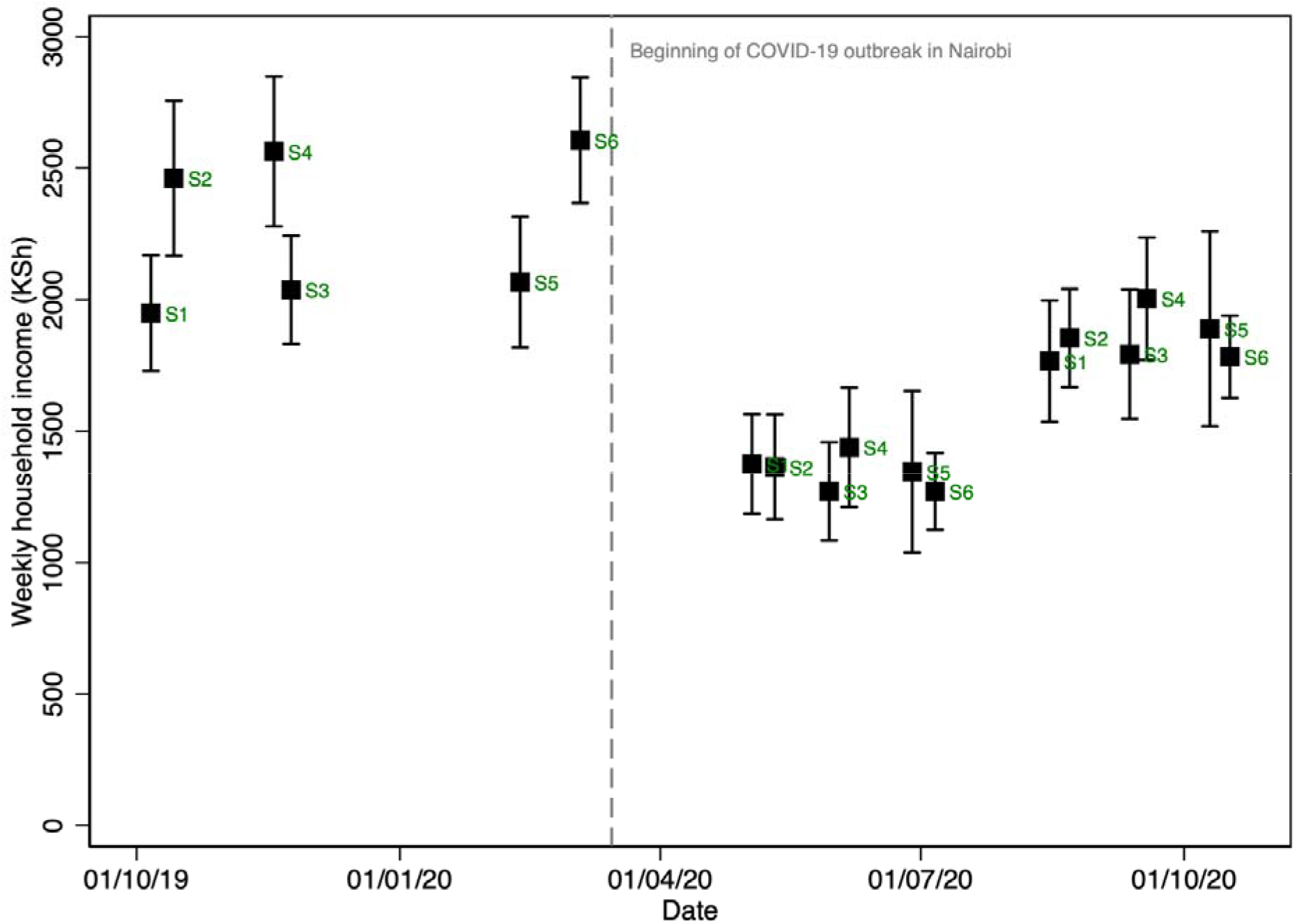
Weekly household income in Kenyan Shillings (KSh) by study site This figure shows self-reported weekly household income in Kenyan Shillings (KSh) over time. Data shown are from a panel of households surveyed both before and after the COVID-19 pandemic. Each point estimate represents data from one of the six sites distributed across Kibera with the vertical bars representing 95% confidence intervals. Sampling weights were applied to account for stratification. Standard errors are clustered by primary sampling unit (multi-household structure). S1 = Site 1; S2 = Site 2; S3 = Site 3; S4 = Site 4; S5 = Site 5; S6= Site 6.

During the pandemic period, the most commonly-cited expenses for which household incomes were insufficient were rent and food. Additionally, 27% of households reported their income was insufficient for healthcare expenses and 46% reported their income was insufficient to obtain drinking water (**Figure 3**). Households attributed their reduced income to fewer labor market opportunities or lower demand for services during the pandemic (43%, the most commonly cited reason), rather than to the direct impacts of the lockdown on their own mobility (8.6%) or the direct health impacts of the pandemic (1.2%) (**Figure 4**).

**Figure 3.**
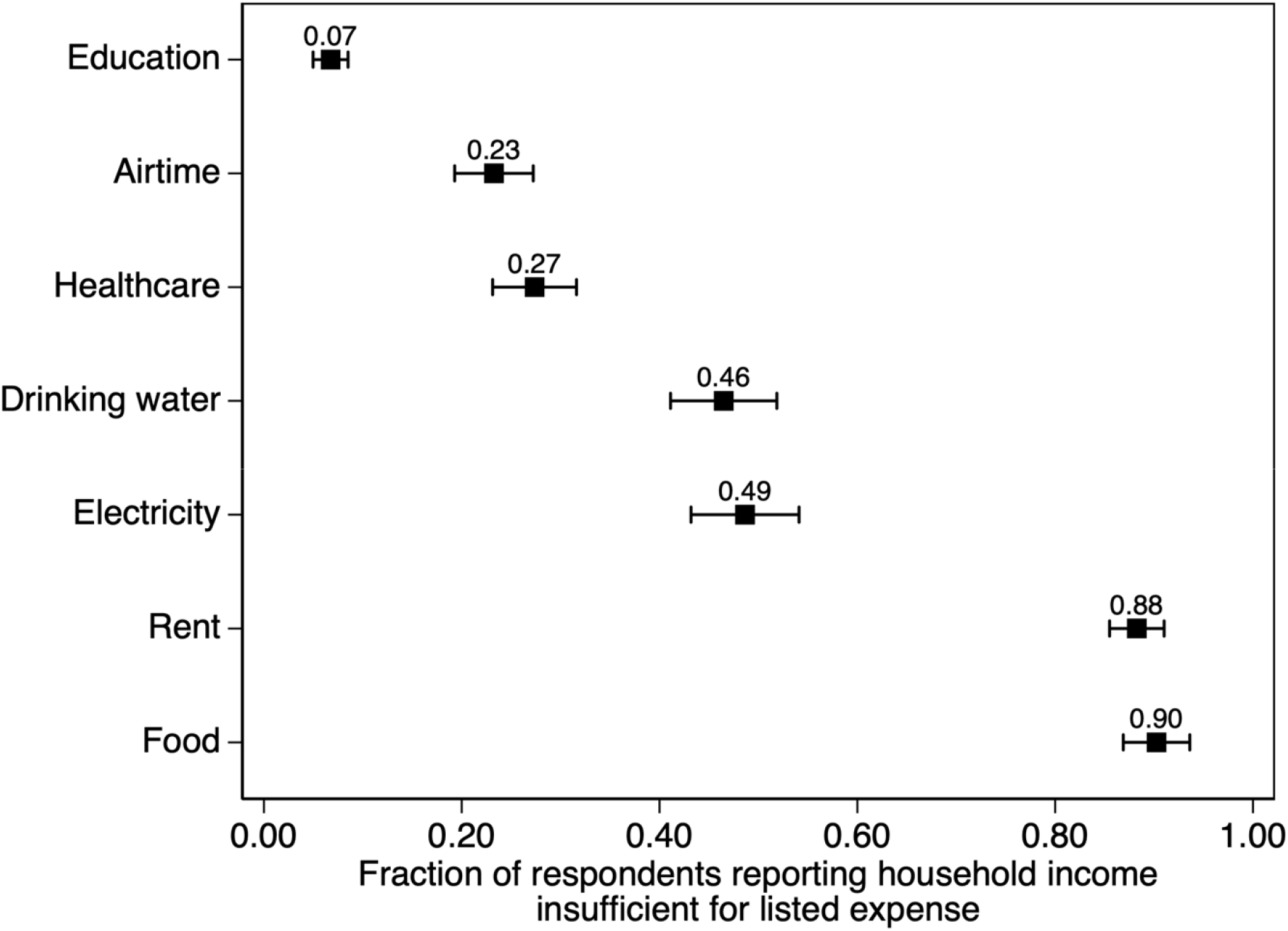
Household expenses for which income is insufficient This figure shows the fraction of all survey respondents reporting that their household income was insufficient for the listed expense between April and October 2020. Respondents could list multiple expenses for which income is insufficient. Horizontal bars represent 95% confidence intervals. Sampling weights were applied to account for stratification. Standard errors are clustered by primary sampling unit (multi-household structure).

**Figure 4.**
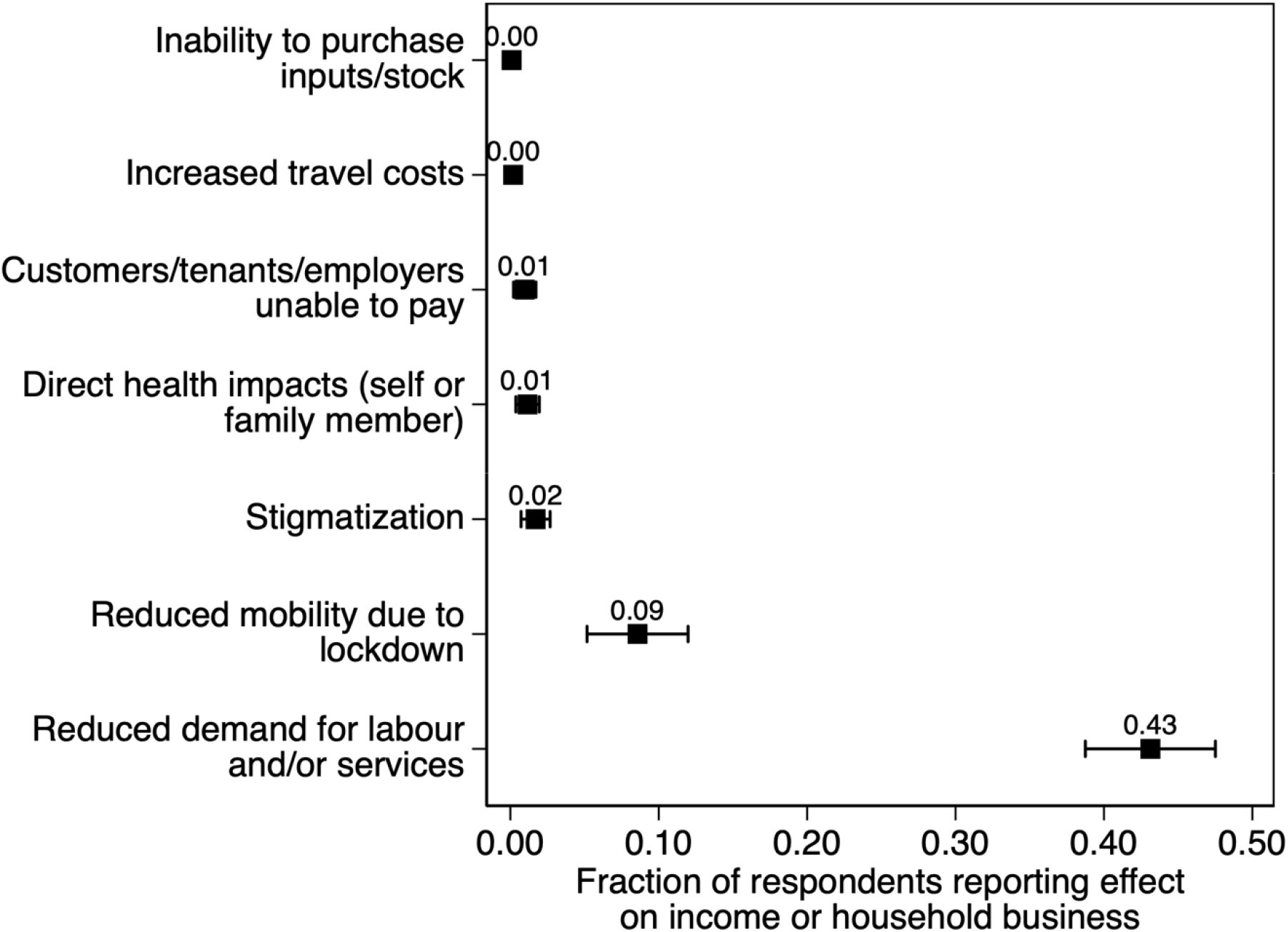
Channels for effects on household income This figure shows the fraction of all survey respondents reporting each of the listed effects on their household income or a business owned by a household member between April and October 2020. Respondents could list multiple effects. Horizontal bars represent 95% confidence intervals. Sampling weights were applied to account for stratification. Standard errors are clustered by primary sampling unit (multi-household structure).

Respondents also reported limited ability to adopt protective behaviors during the pandemic. Nearly all respondents (88% between April and July 2020, rising to 95% between September and October 2020) reported needing to access congregant settings to obtain food or water or for sanitation. The share of individuals reporting at least one symptom among fever, a dry cough, shortness of breath, or anosmia at the time of survey averaged 5% between April and October.

### Estimated drops in income are unlikely to be explained by secular trends, seasonal variation, or non-random loss to follow-up

The estimated decline in household income was unchanged across sensitivity analyses accounting for secular trends, seasonal variation, and non-random loss-to-follow up. Before the COVID-19 pandemic, mean household income was stable or increasing, suggesting that the decline in income during the pandemic was not driven by pre-existing trends (**Supplemental Figures 2 and 3**). The estimated decline in income during the pandemic was essentially unchanged when adjusting for seasonal variation in income using season and quarter-of-year fixed effects (**Supplemental Figure 3**). Our main result, which included movers, was unchanged after excluding movers from the analysis and when limiting the sample to a fully balanced panel that only included households that were successfully surveyed in every wave (**Supplemental Figure 4**). Finally, although response rates were high, the estimated change in incomes could be biased if those respondents who could not be interviewed by phone during the pandemic experienced larger or smaller changes in income than those who were interviewed.

Reassuringly, there was no correlation between the number of times enumerators had to call respondents in order to secure an interview between April and July 2020 and the respondent’s change in household income during versus before the pandemic. If respondents who could not be reached were similar to respondents who were most difficult to reach, this suggests non-random loss-to-follow up is unlikely to explain the observed decline in income during the first months of the pandemic. However, during September and October 2020, respondents who were harder to reach reported a greater recovery in income, possibly because these respondents had returned to the labor force. This pattern suggests that we might underestimate the recovery in income in September and October 2020 (**Supplemental Figure 5**).

## Conclusion

In this longitudinal cohort study of residents of informal settlements in Kenya, we observed a 59% reduction in household income during the COVID-19 pandemic that had yet to recover to baseline levels after six months.

Study limitations include that our study sample is representative of the six sites within the informal settlement but not of the whole settlement of Kibera. We note that we were able to achieve relatively high response rates because of intensive long-term engagement in the six communities. Furthermore, the six communities are distributed across Kibera and the change in income was similar in all communities. There was limited scope to further increase response rates during the pandemic conditions; in cases where enumerators could not reach respondents, they made on average 8.5 attempts to do so. Nonetheless, if loss to follow-up was correlated with changes in income, it could potentially introduce bias into our estimated effects on changes in income. Additionally, since no objective measures of income exist for this population, we depended on self-reported measures of income. However, the effects we observe are consistent across different self-reported measures, suggesting that the results are unlikely to be artefacts of a specific measurement process.

Further research should investigate the long-run consequences of the short-term losses in income we observed, with the aim of understanding how different policy choices might mitigate income losses among residents of informal settlements during the ongoing COVID-19 pandemic and future pandemics.

While the longer-term consequences for welfare remain unknown, the scale of the reduction in income we observed raises serious concerns about the well-being of highly vulnerable residents of informal settlements in sub-Saharan Africa during the pandemic.

## Data Availability

Anonymized replication data are available on request.

## Non-author Contributions

None.

## Access to Data and Data Analysis

Dr. Tompsett had full access to all the data in the study and takes responsibility for the integrity of the data and the accuracy of the data analysis.

## References

1. United Nations Statistics Division: Development Data and Outreach Branch. https://unstats.un.org/sdgs/report/2019/goal-11/.

2. Austrian K, Pinchoff J, Tidwell JB, White C, Abuya T, Kangwana B, et al. COVID-19 related knowledge, attitudes, practices and needs of households in informal settlements in Nairobi, Kenya. 2020.

3. Gil D, Domínguez P, Undurraga EA, Valenzuela E. The Socioeconomic Impact of COVID-19 in Urban Informal Settlements. medRxiv. 2021:2021.01.16.21249935.

4. Armand A, Augsburg B, Bancalari A. Coping with COVID-19 in Slums: Evidence from India. International Growth Centre. November, 2020.

5. Revitalising Informal Settlements and their Environments. The impact of COVID-19 in Fijian informal settlements: Results of a rapid phone survey conducted in May 2020. June, 2020.

6. Quaife M, Van Zandvoort K, Gimma A, Shah K, McCreesh N, Prem K, et al. The impact of COVID-19 control measures on social contacts and transmission in Kenyan informal settlements. BMC medicine. 2020;18(1):1–11.

7. Corburn J, Vlahov D, Mberu B, Riley L, Caiaffa WT, Rashid SF, et al. Slum health: arresting COVID-19 and improving well-being in urban informal settlements. Journal of urban health. 2020;97(3):348–57.

8. Egger D, Miguel E, Warren SS, Shenoy A, Collins E, Karlan D, Parkerson D, Mobarak AM, Fink G, Udry C, Walker M. Falling living standards during the COVID-19 crisis: Quantitative evidence from nine developing countries. Science Advances. 2021 Feb 1;7(6):eabe0997.

9. Malani A, Shah D, Kang G, Lobo GN, Shastri J, Mohanan M, Jain R, Agrawal S, Juneja S, Imad S, Kolthur-Seetharam U. Seroprevalence of SARS-CoV-2 in slums versus non-slums in Mumbai, India. The Lancet Global Health. 2021 Feb 1;9(2):e110–1.

10. Mukeku J. Urban Slum Morphology and Socio-economic Analogies: A Case Study of Kibera Slum, Nairobi, Kenya. Urbanisation. 2018 May;3(1):17–32.

11. Uyoga S, Adetifa IM, Karanja HK, Nyagwange J, Tuju J, Wanjiku P, Aman R, Mwangangi M, Amoth P, Kasera K, Rombo C. Seroprevalence of anti–SARS-CoV-2 IgG antibodies in Kenyan blood donors. Science. 2021 Jan 1;371(6524):79–82.

12. Makayotto L, Oluga O, Osoro E. Findings of Covid-19 Antibody Survey in Nairobi City County Conducted in November 2020. Policy brief. January 26, 2021.

