## Supplemental Materials for "Changes to household income in a Kenyan informal settlement during COVID-19"

**Supplemental Figure 1.** Fraction of respondents reporting household income sufficient for household needs.

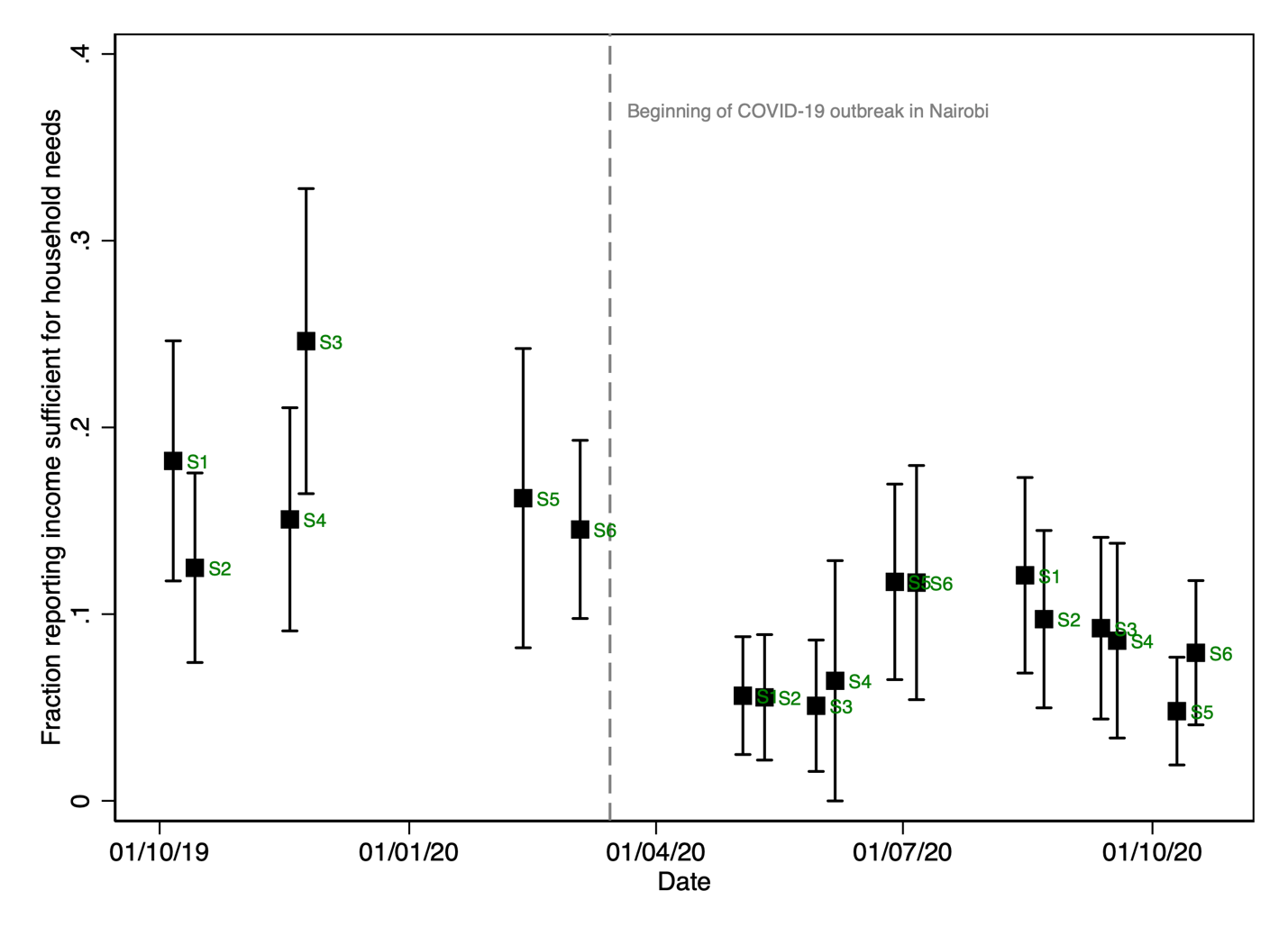

This figure shows the fraction of respondents reporting that their household income is sufficient for their households needs over time. Data shown are from a panel of households surveyed both before and after the COVID-19 pandemic. Each point estimate represents data from one of the six sites distributed across Kibera with the vertical bars representing 95% confidence intervals. Sampling weights were applied to account for stratification. Standard errors are clustered by primary sampling unit (multi-household structure). S1 = Site 1; S2 = Site 2; S3 = Site 3; S4 = Site 4; S5 = Site 5; S6= Site 6.

**Supplemental Figure 2.** Weekly household income in Kenyan Shillings (KSh) by study site in sites with two pre-COVID waves of data.

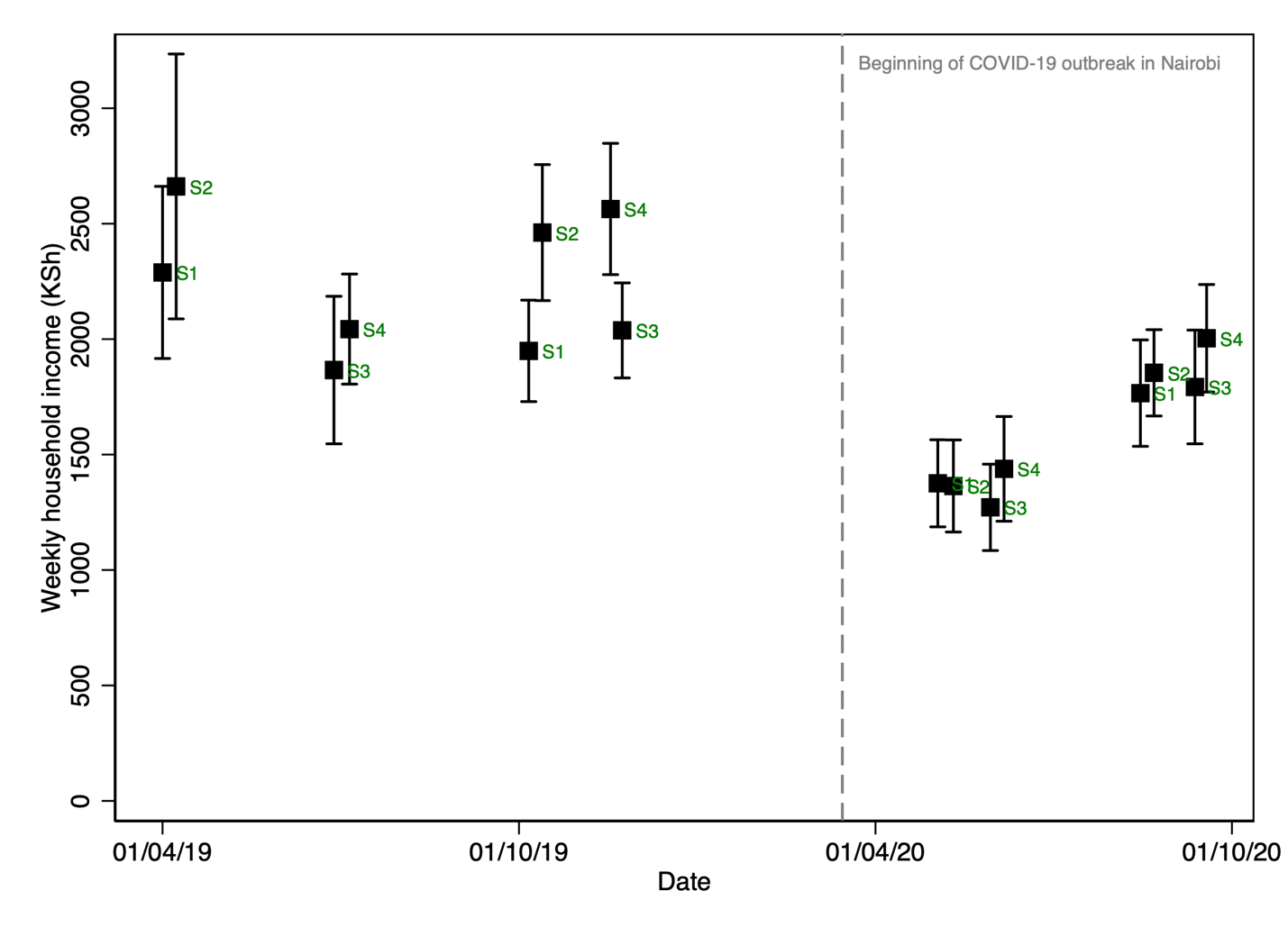

This figure shows self-reported weekly household income in Kenyan Shillings (KSh) over time. Data shown are from a balanced panel of households surveyed both before and after the COVID-19 pandemic. We restrict to a balanced panel to separate secular trends from sample changes but in practice the results are insensitive to different sample restrictions. Each point estimate represents data from one of the four sites for which we have two pre-COVID rounds of data with the vertical bars representing 95% confidence intervals. Sampling weights were applied to account for stratification. Standard errors are clustered by primary sampling unit (multi-household structure). S1 = Site 1; S2 = Site 2; S3 = Site 3; S4 = Site 4.

**Supplemental Figure 3.** Robustness to controlling for seasonality.

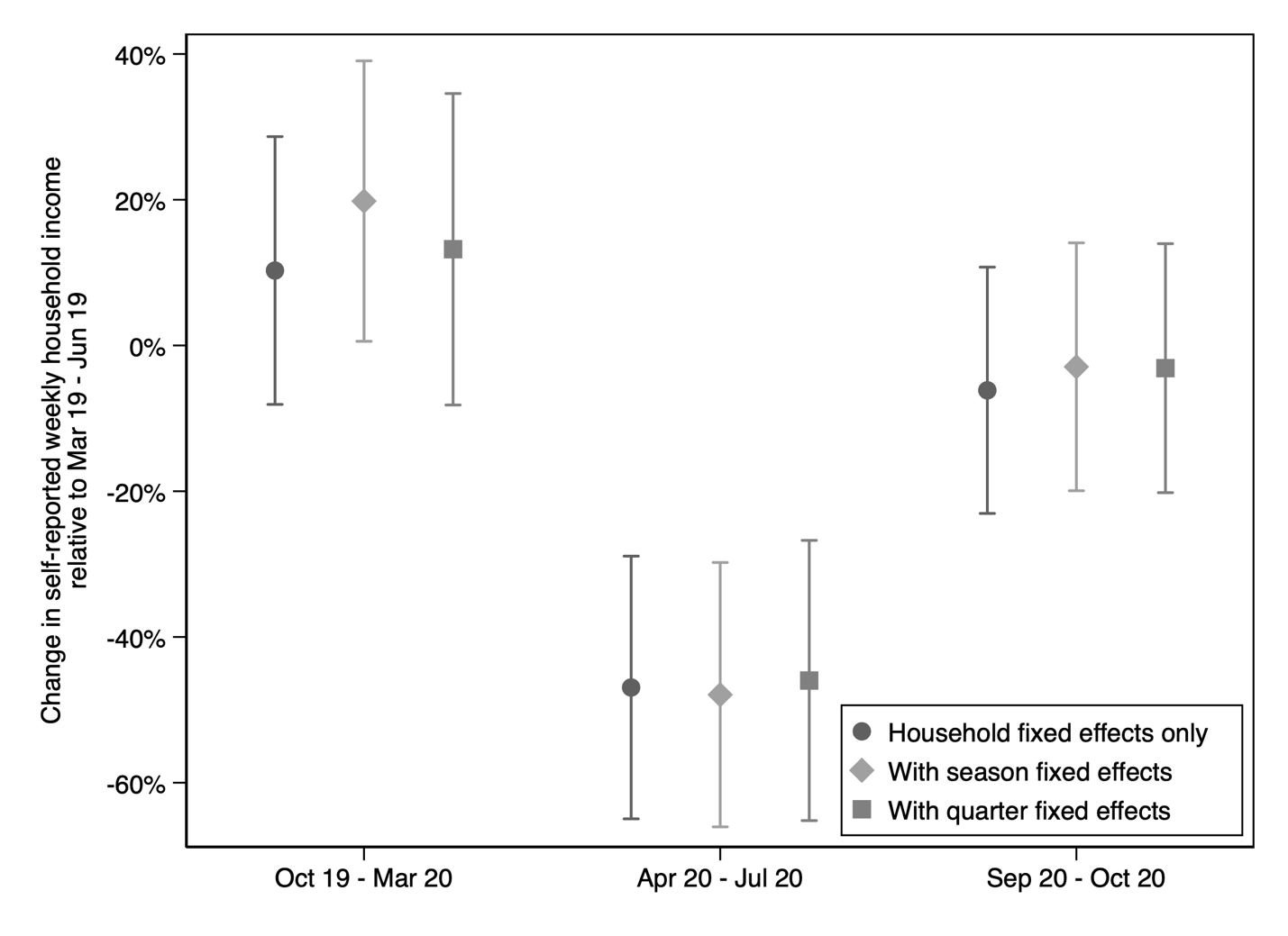

This figure shows estimated change in weekly household income in Kenyan Shillings (KSh) relative to the baseline period (March-June 2019). Data shown are from a balanced panel of households surveyed both before and after the COVID-19 pandemic. The vertical bars representing 95% confidence intervals. Sampling weights were applied to account for stratification. Standard errors are clustered by primary sampling unit (multi-household structure). Season fixed effects absorb variation by season of year (two rain seasons, two dry seasons). Quarter fixed effects absorb variation by quarter of year.

**Supplemental Figure 4.** Robustness to changes in sample.

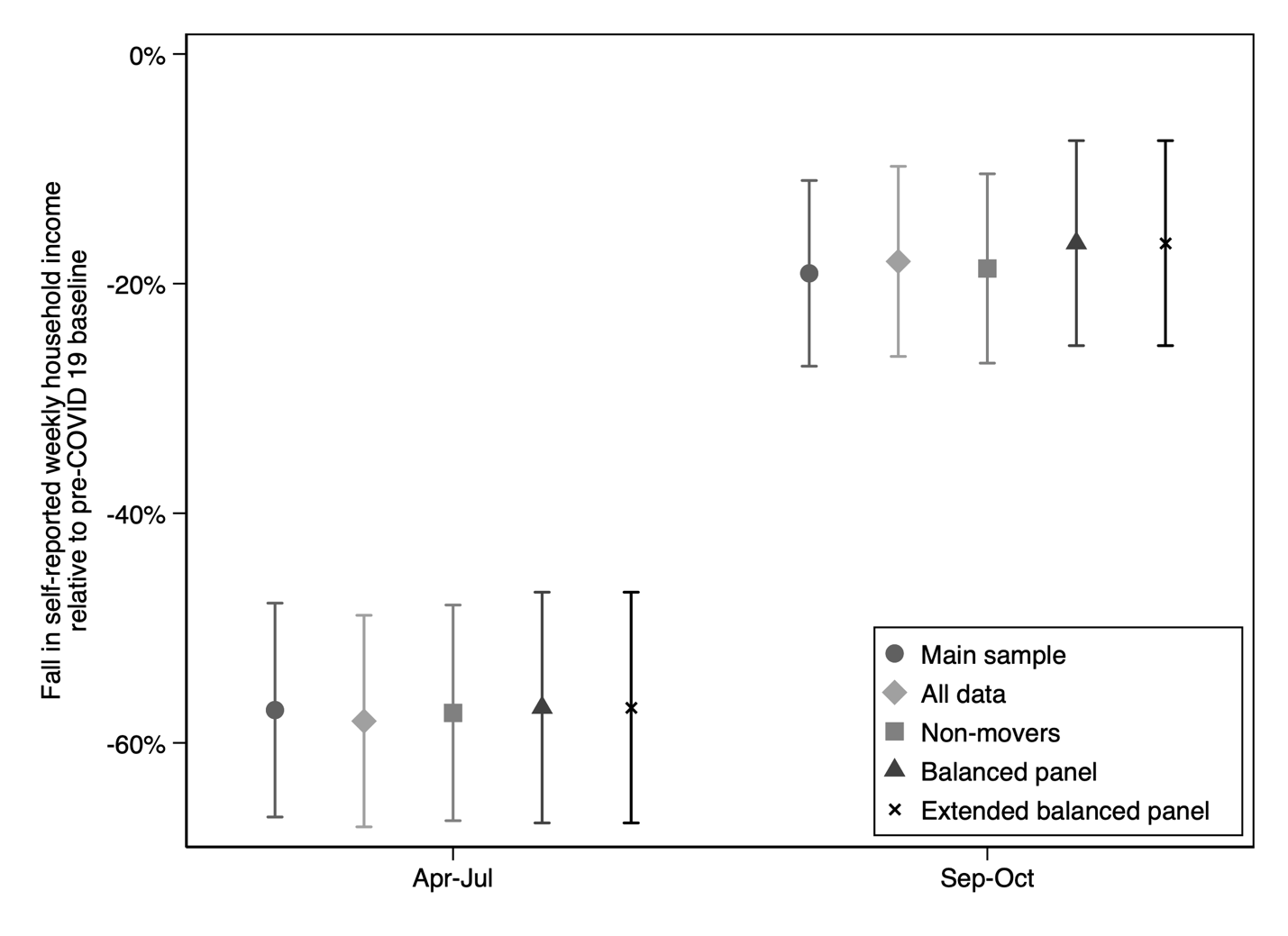

This figure shows estimated change in weekly household income in Kenyan Shillings (KSh) relative to the baseline period (March-June 2019). Data shown are from a panel of households surveyed both before and after the COVID-19 pandemic. All estimates account for pre-pandemic differences between households in income using household fixed effects. The vertical bars representing 95% confidence intervals. Sampling weights were applied to account for stratification. Standard errors are clustered by primary sampling unit (multi-household structure). Main sample consists of respondents who were surveyed at least one before and after the pandemic. All data includes all respondents surveyed at least once during the entire study period. Non-movers excludes respondents who leave Kibera during the study period from the main sample. The balanced panel includes only respondents with non-missing data from all three waves. The extended balanced panel includes respondents with non-missing data from four waves, including two pre-pandemic waves.

**Supplemental Figure 5.** Correlation between change in income relative to pre-pandemic period and number of attempts required to successfully complete interview

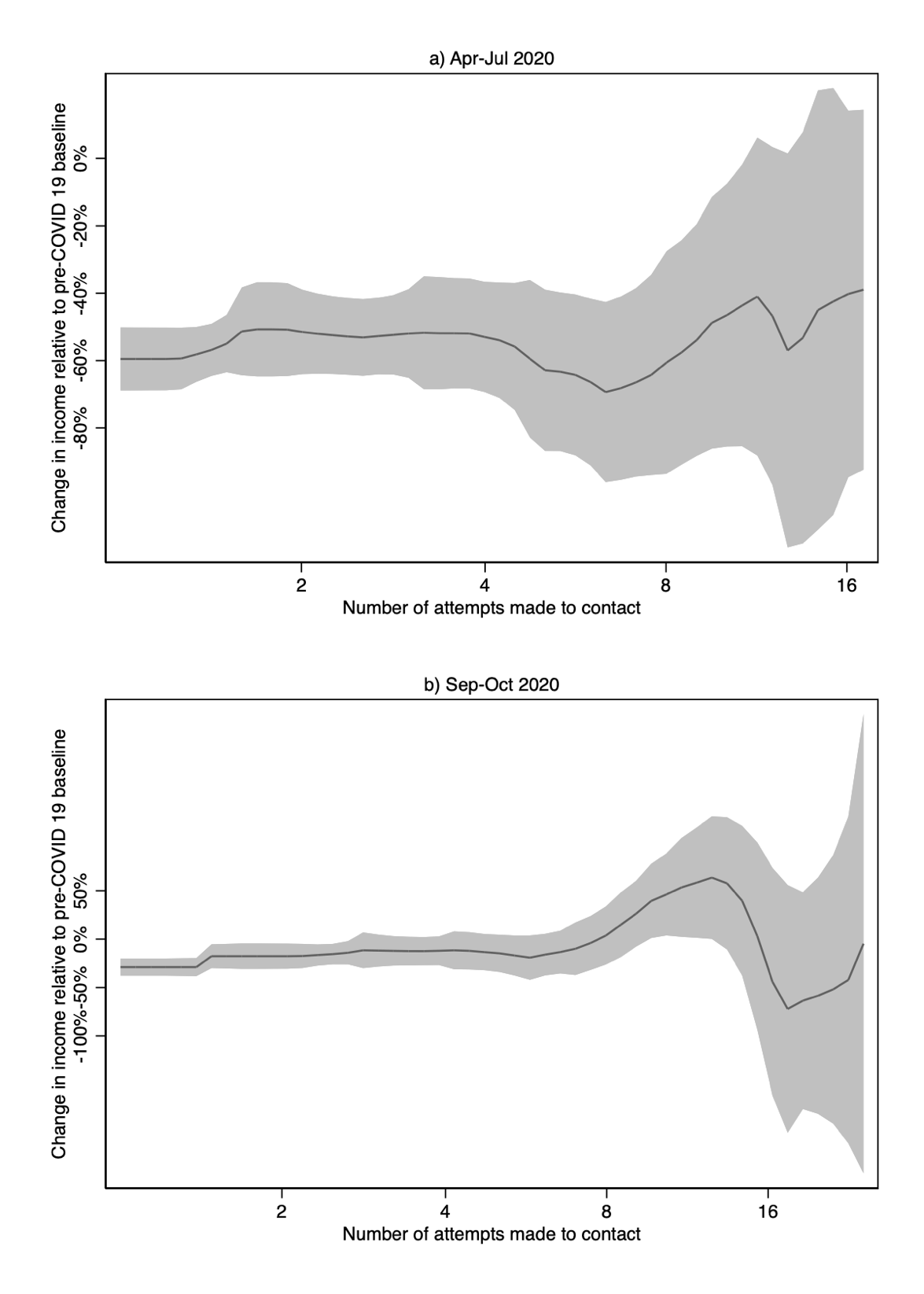

This figure plots the relationship between estimated change in weekly household income in Kenyan Shillings (KSh) relative to the baseline period (March-June 2019) and log number of attempts needed to secure an interview. Graph plots results of a local linear regression. The shaded area shows 95% confidence intervals.
